# Real-world Experience with Favipiravir for Treatment of COVID-19 in Thailand: Results from a Multicenter Observational Study

**DOI:** 10.1101/2020.06.24.20133249

**Authors:** Pinyo Rattanaumpawan, Supunnee Jirajariyavej, Kanokorn Lerdlamyong, Nattawan Palavutitotai, Jatuporn Saiyarin

**Author notes:** **Correspondence to:** Pinyo Rattanaumpawan, MD, MSCE, PhD, 2 Wanglang Road, Bangkoknoi, Bangkok 10700, Thailand.

## Abstract

**Background:** Favipiravir is a broad-spectrum oral antiviral agent that shows in vitro activity against SARS-CoV-2. Presently, data on the effectiveness and optimal dosage of favipiravir for treating COVID-19 is limited.

**Methods:** We conducted a retrospective observational study of hospitalized adult patients with COVID-19 at five tertiary care hospitals in Thailand. We reviewed patient charts to obtain all necessary data.

**Results:** Among 247 COVID-19 patients, 63 (23.0%) received ≥1 dose of favipiravir. Of these, 27.0% required an O_2_-nasal cannula, 9.5% required non-invasive ventilation and/or high-flow O_2_-therapy, and 6.4% required invasive mechanical ventilation and/or ECMO. The median baseline NEWS2 score was 5(0–16). The Day-7 clinical improvement rate [95%CI] was 66.7%[53.7–78.0%] in all patients, 92.5%[75.7%–99.1%] in patients who did not require O_2_-supplementation, and 47.2%[0.4%–64.5%] in patients who required O_2_-supplementation. No life-threatening adverse events were identified. The 28-day mortality rate was 4.8%. Multivariate analysis revealed three poor prognostic factors for Day-7 clinical improvement [odds ratio (95%CI); *p*-value]: older age [0.94 (0.89–0.99); *p*=0.04], higher baseline NEWS2 score [0.64 (0.47–0.88); *p*=0.006], and lower favipiravir loading dose (≤45 mg/kg/day) [0.04 (0.005–0.4); *p*=0.006].

**Conclusions:** Our study reports the promising effectiveness of favipiravir for treating COVID-19 patients. In addition to older age and a high baseline NEWS2 score, a low loading dose of favipiravir (≤45 mg/kg/day) was also identified as a poor prognostic factor for early clinical improvement. Further studies to explore the optimal dose and the optimal timing of drug initiation for favipiravir should be performed.

## INTRODUCTION

As of 16-Jun-2020, a total of 7,941,791 COVID-19 cases with 434,796 deaths have been reported globally.^1^ This pandemic disease is caused by a novel coronavirus, named severe acute respiratory syndrome coronavirus 2 (SARS-CoV-2). SARS-CoV-2 is a single-stranded RNA beta-coronavirus encoding an RNA-dependent RNA polymerase (RdRp) and proteases. Both RdRp and viral proteases are considered important targets for potentially therapeutic agents. Hundreds of clinical studies are actively investigating a variety of promising agents (e.g., remdesivir, favipiravir, lopinavir, hydroxychloroquine, and interferon-alpha)^2^; however, data on the efficacy of these potentially therapeutic agents are still limited.

Favipiravir, a purine nucleic acid analog, is a broad-spectrum oral antiviral agent that inhibits the *RdRp* of RNA viruses.^3^ This agent shows in vitro activity against many RNA viruses, including arenaviruses, bunyaviruses, flaviviruses, Ebola virus, and influenza virus, as well as SARS-CoV-2.^4, 5^ Currently, two registered clinical studies of favipiravir among COVID-19 patients have already reported their results.^6^ The first study (ChiCTR2000029600) was a small, open-label, nonrandomized control study of 80 patients with COVID-19 conducted to compare the efficacy of favipiravir plus aerosolized interferon-alpha with that of lopinavir/ritonavir plus aerosolized interferon-alpha. In that study, the favipiravir group showed a significantly shorter time to viral clearance and a significantly higher improvement rate in chest imaging, after adjustment for potential confounders.^7^ The second study (ChiCTR200030254) was a randomized control trial (RCT) of 240 patients with COVID-19 pneumonia. That study, currently available as a preprint on MedRxIV, reported a significantly higher clinical recovery rate on Day 7 in the favipiravir group compared with the arbidol group (71.43% vs. 55.86%; *p* = 0.02).^8^

In February 2020, favipiravir was made available for use in Thailand under emergency procurement by the Department of Disease Control of Thailand. In early March 2020, Thailand national clinical practice guidelines (CPG) for COVID-19 management recommended the initiation of favipiravir therapy in only patients with severe COVID-19 pneumonia (pneumonia with required high-flow O_2_-supplementation, non-invasive mechanical ventilation, or invasive mechanical ventilation to maintain a patient O_2_-saturation of 90% or more). In early May 2020, the national CPG were revised, and favipiravir is now also recommended for the treatment of mild pneumonia (i.e., abnormal chest-x-ray without desaturation). The benefits of early favipiravir initiation in mild COVID-19 cases are in need of further investigation.

The standard dose of favipiravir for treating influenza infection is 1600 mg twice daily on Day 1, followed by 600 mg twice daily on Days 2–5.^9^ A maximal loading dose of 3000 mg twice daily on Day 1 and a maintenance dose of 1200 mg twice daily on Days 2–9 were safely used in a previous Ebola study.^10^ Given that the optimal dose of favipiravir for treating COVID-19 is still uncertain, the Thailand national CPG recommend a fixed loading dose of 1600 mg twice daily on Day 1, followed by 600 mg twice daily on Days 2–10. A higher loading dose (60 mg/kg/day, MKD) and maintenance dose (20 MKD) are recommended in patients with a body mass index (BMI) of ≥35.

Presently, data on the effectiveness and optimal dosage of favipiravir for treating COVID-19 is limited. Therefore, we conducted a retrospective study to explore these issues.

## MATERIALS AND METHODS

### Study Design

We conducted a retrospective observational study of COVID-19 patients who were hospitalized at any of five tertiary care hospitals in Thailand (i.e., Siriraj, Taksin, Vachira Phuket, Lerdsin, and Central hospitals) during 1 January – 30 April 2020. The study protocol was approved with a waiver of informed consent by the institutional review boards of all involved hospitals.

### Inclusion and Exclusion Criteria

We enrolled all hospitalized patients aged at least 18 years who had reverse transcription PCR-confirmed SARS-CoV-2 based on a respiratory specimen (nasopharyngeal, oropharyngeal, sputum, endotracheal aspirate, or bronchoalveolar lavage sample) and received at least one dose of favipiravir. Patients who expired or were discharged from the hospital within 24 hours after hospitalization were excluded.

### Data Collection and Study Definition

We reviewed patient charts to obtain all necessary data, including demographic data, clinical data, laboratory data, and the hospital stay length. We also recorded the daily National Early Warning Score 2 (NEWS2 score). Details regarding the NEWS2 score have been published elsewhere.^11^ The primary outcome was the rate of clinical improvement within seven days of favipiravir therapy (Day-7 clinical improvement), and the secondary outcomes were the Day-14 and Day-28 clinical improvement rates.

Clinical improvement was defined as a one-point reduction in baseline status (on the first day of favipiravir therapy) on a six-point disease severity scale at the time of evaluation. The six-point disease severity scale was categorized as follows: 6-death; 5-hospitalization for extracorporeal membrane oxygenation (ECMO) or mechanical ventilation; 4-hospitalization for non-invasive ventilation or high-flow O_2_-therapy; 3-hospitalization for supplemental O_2_; 2-hospitalization without the need for O_2_-supplementation but requiring ongoing medical care; and 1-discharge or normalization of all vital signs and saturation of peripheral O_2_ of >94% on room air for at least 24 hours.

### Statistical Analysis

Categorical variables are summarized by frequency and percentage, whereas continuous variables are summarized by the median and range. Univariate analyses were performed using the Fisher-exact test for categorical data. The Mann-Whitney U test was used for continuous data. To identify the factors independently associated with the Day-7 clinical improvement, we performed a subsequent multivariate analysis including all potentially significant variables with a *p*-value of ≤0.20 in a stepwise fashion.

For all calculations, a two-tailed *p*-value of <0.05 was considered statistically significant. All calculations were performed using STATA version 14.1 (Stata Corp, College Station TX).

## RESULTS

During the study period, there were a total of 274 COVID-19 patients hospitalized in the participating hospitals, of which 63 patients (23.0%) received favipiravir. The baseline demographics and characteristics of all patients are listed in Table 1.

**Table 1.** Baseline demographics and characteristics of all patients.

| Variables | All (n = 63) | Day 7 Clinical improvement |  | p-value |
| --- | --- | --- | --- | --- |
|  |  | Yes (n = 42) | No (n = 21) |  |
| Age, median (range), year | 48 (22–85) | 47 (23–72) | 59 (22–85) | 0.02 |
| Male sex | 39 (61.9%) | 25 (59.5%) | 14 (66.7%) | 0.78 |
| Body weight, median (range), kg | 69 (45–125) | 68 (51–125) | 76 (45–120) | 0.08 |
| Body mass index median (range), kg/m<sup>2</sup> | 26.1 (19.0–43.8) | 25.0 (19.0–43.8) | 27.9 (20.8–39.2) | 0.04 |
| **Duration between, median (range), day** |  |  |  |  |
| Symptom onset and admission date | 6 (0–28) | 6 (0–28) | 8 (0–15) | 0.08 |
| Admission date and Day-1 of favipiravir therapy | 1 (-8–10) | 1 (-3–10) | 0 (-8–5) | 0.002 |
| Symptom onset and Day-1 of favipiravir therapy | 8 (0–28) | 8 (2–28) | 8 (0–11) | 0.60 |
| **Exposure risk** |  |  |  |  |
| Contact with confirmed COVID-19 cases | 26 (41.3%) | 19 (45.2%) | 7 (33.3%) | 0.42 |
| Travel abroad | 7 (11.1%) | 5 (11.2%) | 2 (9.5%) | 1.00 |
| Contact with a foreigner | 11 (17.5%) | 8 (19.1%) | 3 (14.3%) | 0.74 |
| Travel to a local area with clustered cases | 38 (60.3%) | 28 (66.7%) | 10 (47.6%) | 0.18 |
| **Underlying diseases** |  |  |  |  |
| Heart disease and hypertension | 9 (14.3%) | 7 (16.7%) | 2 (9.5%) | 0.71 |
| Diabetes mellitus | 17 (27.0%) | 11 (26.2%) | 6 (28.6%) | 1.00 |
| Chronic lung disease | 4 (6.4%) | 2 (7.1%) | 1 (4.8%) | 1.00 |
| Chronic kidney disease | 4 (6.4%) | 3 (7.1%) | 1 (4.8%) | 1.00 |
| Chronic liver disease | 3 (4.8%) | 3 (7.1%) | 0 (0%) | 0.55 |
| Solid cancer | 4 (6.4%) | 2 (7.1%) | 1 (4.8%) | 1.00 |
| Others | 4 (6.4%) | 2 (7.1%) | 2 (9.5%) | 0.60 |
| **Clinical presentation upon admission** |  |  |  |  |
| Fever or body temperature of >37.5 °C | 55 (87.3%) | 36 (85.7%) | 19 (90.5%) | 0.71 |
| Sore throat | 44 (69.8%) | 27 (64.3%) | 17 (81.0%) | 0.25 |
| Rhinorrhea | 16 (25.4%) | 13 (31.0%) | 3 (14.3%) | 0.22 |
| Cough | 47 (74.6%) | 30 (71.4%) | 17 (81.0%) | 0.54 |
| Headache | 11 (17.5%) | 8 (19.1%) | 3 (14.3%) | 0.74 |
| Myalgia | 17 (27.0%) | 12 (28.6%) | 5 (23.8%) | 0.77 |
| Diarrhea | 8 (12.7%) | 6 (14.3%) | 2 (9.5%) | 0.71 |
| Shortness of breath | 27 (42.9%) | 14 (33.3%) | 13 (61.9%) | 0.06 |
| Illness severity at the time of favipiravir initiation |  |  |  |  |
| NEWS2 score, median (range) | 5 (0–16) | 4 (0–11) | 5 (0–16) | 0.003 |
| Six-point disease severity scale, median (range) | 2.5 (1–5) | 2 (1–4) | 3 (2–5) | <0.001 |
| 1 – No O<sub>2</sub>-supplementation with O<sub>2</sub>-saturation of >94% | 4 (6.4%) | 4 (6.4%) | 0 (0) | <0.001 |
| 2 – No O<sub>2</sub>-supplementation with O<sub>2</sub>-saturation of ≤94% | 23 (36.4%) | 21 (50.0%) | 2 (9.5%) |  |
| 3 –Requiring O<sub>2</sub>-supplementation | 28 (44.4%) | 16 (40.1%) | 12 (57.1%) |  |
| 4 –Requiring high-flow O<sub>2</sub>-supplementation or non-invasive mechanical ventilation | 4 (6.4%) | 1 (2.4%) | 3 (14.3%) |  |
| 5 –Requiring invasive mechanical ventilation and/or extracorporeal membrane oxygenation | 4 (6.4%) | 0 (0%) | 4 (19.1%) |  |
| Baseline laboratory values* |  |  |  |  |
| Hemoglobin, median (range), (mg/dl) | 14.0 (8.0–18.0) | 14.0 (9.0–17.0) | 13.5 (8.0–18.0) | 0.48 |
| White blood cell count, median (range), (cell/mm<sup>3</sup>) | 5735<br>(2910–41300) | 5420<br>(2910–41300) | 6810<br>(3180–15750) | 0.03 |
| Serum creatinine, median (range), (mg/dl) | 0.9 (0.3–22.9)<br>(n = 58) | 0.9 (0.4–22.9)<br>(n = 27) | 0.9 (0.33–5.1)<br>(n = 21) | 0.67 |
| Serum albumin, median (range), (mg/dl) | 4.0 (1.8–4.9)<br>(n = 53) | 4.2 (1.8–5.0)<br>(n = 33) | 3.5 (2.6–4.1)<br>(n = 20) | 0.002 |
| Serum lactate dehydrogenase, median (range), (mg/dl) | 404 (145–1094)<br>(n = 30) | 382 (145–567)<br>(n = 17) | 453 (313–1094)<br>(n = 13) | 0.03 |
| **Indication of favipiravir therapy** |  |  |  |  |
| Abnormal chest imagining only | 26 (41.3%) | 24 (57.1%) | 2 (9.5%) | <0.001 |
| Required O<sub>2</sub>-supplementation only | 3 (4.7%) | 2 (4.7%) | 1 (4.8%) |  |
| Abnormal chest imaging and required O<sub>2</sub>-supplementation | 34 (54.0%) | 16 (38.1%) | 18 (85.7%) |  |
| **Favipiravir regimen** |  |  |  |  |
| Dose per bw, median (range), mg/kg/day |  |  |  |  |
| Loading dose | 47.4 (29.1–71.1) | 49.2 (29.1–62.7) | 45.7 (29.6–71.1) | 0.47 |
| Maintenance dose | 17.9 (10.9–26.7) | 18.5 (10.9–23.5) | 17.1 (11.1–26.7) | 0.37 |
| Potentially sub-therapeutic dose |  |  |  |  |
| Loading dose of ≤45 MKD | 21 (33.3%) | 11 (26.2%) | 10 (47.6%) | 0.10 |
| Maintenance dose of ≤15 MKD | 48 (76.2%) | 33 (78.6%) | 15 (71.4%) | 0.55 |
| Duration of therapy, median (range), day | 12 (2–17) | 11.5 (2–16) | 12 (2–17) | 0.02 |
| **Other medications used\*\*** |  |  |  |  |
| Any chloroquine-based agent | 62 (98.4%) | 41 (97.6%) | 21 (100%) | 1.00 |
| Hydroxychloroquine | 54 (85.7%) | 36 (85.7%) | 18 (85.7%) | 1.00 |
| Chloroquine | 14 (22.2%) | 8 (19.1%) | 6 (28.6%) | 0.52 |
| Any protease inhibitor | 61 (96.8%) | 40 (95.2%) | 21 (100,0%) | 0.55 |
| Darunavir/ritonavir | 51 (81.0%) | 35 (83.3%) | 16 (76.2%) | 0.51 |
| Lopinavir/ritonavir | 22 (34.9%) | 13 (31.0%) | 9 (42.9%) | 0.26 |
| Azithromycin | 31 (49.2%) | 17 (40.5%) | 14 (66.7%) | 0.06 |
| Steroid | 8 (12.7%) | 5 (11.9%) | 3 (14.3%) | 1.00 |
| Tocilizumab | 4 (6.4%) | 1 (2.4%) | 3 (14.3%) | 0.10 |

**Note.** \*Earliest results of a test obtained within the first 7 days of admission (missing data was replaced by the mean value of the variable)
\*\* Medications used within 2 days before or after the initiation of favipiravir therapy

The median age of favipiravir-treated COVID-19 patients was 48 (22–85) years, and 39 of these patients (61.9%) were male. Most patients had fever (87.3%), sore throat (69.8%), or cough (74.6%) as the clinical presentation. The median duration between the symptom onset and the admission date was 6 (0–28) days, while the median duration between the symptom onset and the first day of favipiravir therapy was 8 (0–28) days.

At baseline (Day 1 of favipiravir therapy), 17 patients (27.0%) required O_2_-supplementation via nasal cannula, 6 patients (9.5%) required non-invasive ventilation and/or high-flow O_2_-therapy, and 4 patients (6.4%) required invasive mechanical ventilation and/or ECMO, while the remainder did not required O_2_-supplementation. The median baseline NEWS2 score was 5 (0–16).

The median loading dose of favipiravir was 47.4 (29.1–71.1) MKD, and one-third of enrolled patients (33.3%) received a loading dose of ≤45 MKD. The median maintenance dose of favipiravir was 17.9 (10.9–26.7) MKD, and 76.2% of the subjects received a maintenance dose of ≤15 MKD. The median duration of favipiravir therapy was 12 (2–17) days. Within two days of initiating favipiravir treatment, nearly all patients were prescribed a chloroquine-based agent (98.4%) and a protease inhibitor (96.8%); half of them also received azithromycin (49.2%). Only a few patients received a steroid (12.7%) or tocilizumab (6.4%) at this time.

### Hospital Course and Treatment Outcomes

Details regarding the hospital course and treatment outcomes are shown in Table 2. The Day-7, Day-14, and Day-28 clinical improvement rates, stratified by the requirement for O_2_-supplementation are depicted in Figure 1. The Day-7 clinical improvement rate [95%CI] was 66.7% [53.7–78.0%] in all patients, 92.5%[75.7–99.1%] in patients who did not require O_2_-supplementation (a six-point disease severity scale score of 1–2), and 47.2% [0.4–64.5%] in patients who required O_2_-supplementation (a six-point severity scale score of 3–5). The Day-14 clinical improvement rates for all patients, those who did not require O_2_-supplementation, and those who required O_2_-supplementation were 85.7% [74.6%–93.2%], 100.0% [87.2%– 1.00%], and 75.0% [57.8%–87.9%], respectively. Nearly all patients who required O_2_-supplementation (96.1%) had clinical improvement within 28 days.

**Table 2.** Hospital course and treatment outcomes

| Variables | All patients (n = 63) |
| --- | --- |
| **Clinical improvement** |  |
| Day-7 clinical improvement | 42 (66.7%) |
| Patients who did not require O<sub>2</sub>-supplementation (n = 27) | 25 (92.6%) |
| Patients who required O<sub>2</sub>-supplementation (n = 36) | 17 (47.2%) |
| Day-14 clinical improvement | 54 (85.7%) |
| Patients who did not require O<sub>2</sub>-supplementation (n = 27) | 27 (100.0%) |
| Patients who required O<sub>2</sub>-supplementation (n = 36) | 27 (75.0%) |
| Day-28 clinical improvement | 57 (90.5%) |
| Patients who did not require O<sub>2</sub>-supplementation (n = 27) | 27 (100.0%) |
| Patients who required O<sub>2</sub>-supplementation (n = 36) | 30 (83.3%) |
| ICU duration, median (range), day | 0 (0–46) |
| Required invasive mechanical ventilation or ECMO* during hospitalization | 8 (12.7%) |
| Before initiation of favipiravir | 4 (6.3%) |
| After initiation of favipiravir | 4 (6.3%) |
| 14-day mortality rate | 1 (1.6%) |
| 28-day mortality rate | 3 (4.8%) |
| In-hospital mortality rate | 5 (7.9%) |
| Length of hospital stay, median (range), day | 15 (2–47) |
| **Adverse drug reactions** | 39 (61.9%) |
| Diarrhea | 34 (54.0%) |
| Hepatitis | 4 (6.4%) |
| QT interval prolongation | 4 (6.4%) |
| Nausea and vomit | 5 (7.9%) |
| Superimposed bacterial infection | 8 (12.7%) |

**Note.** \*ECMO: extracorporeal membrane oxygenation

**Figure 1.**
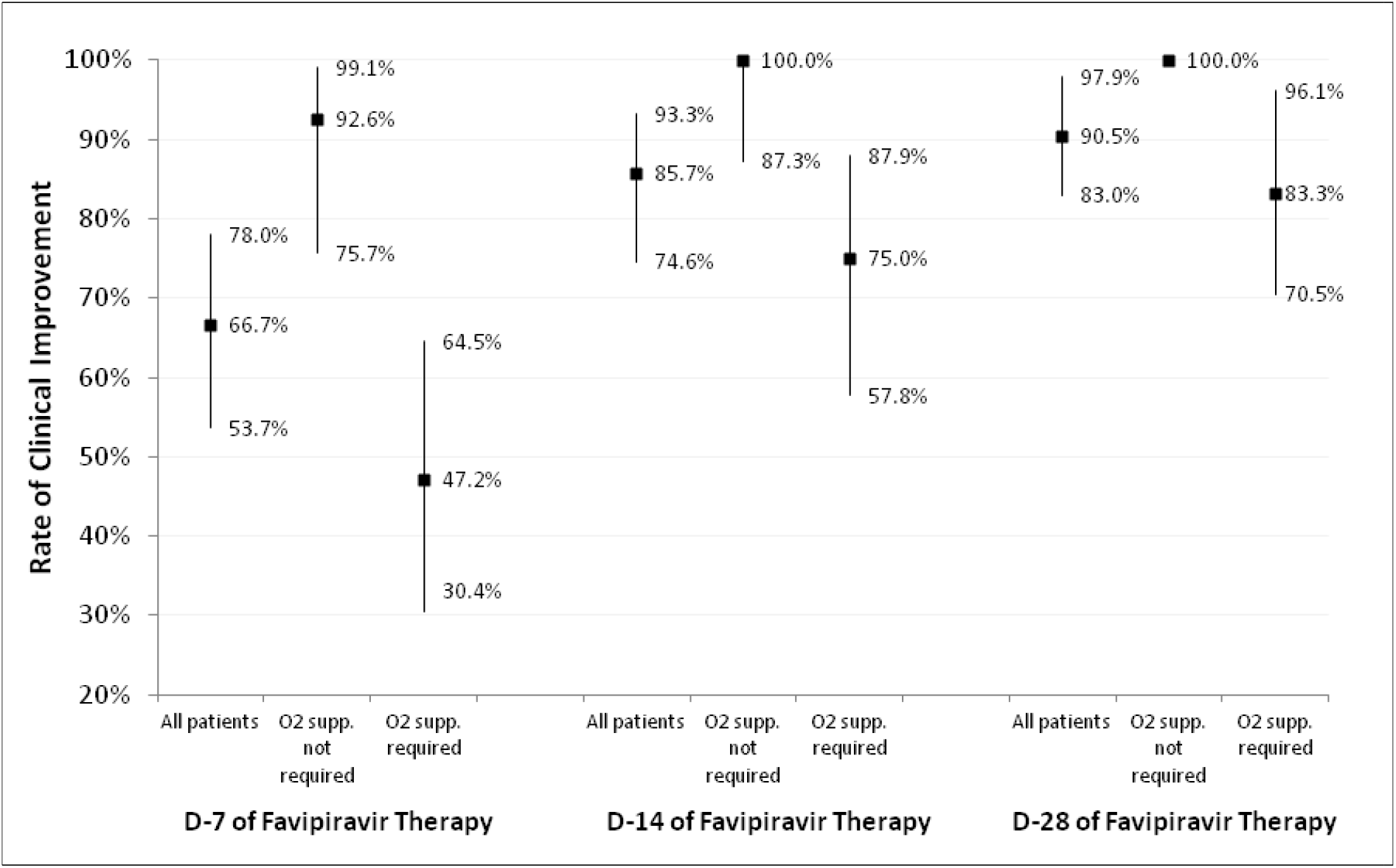
Rate of clinical improvement on Day 7, Day 14, and Day 28 of favipiravir therapy, stratified by the requirement for O_2_-supplementation.

Of the 63 favipiravir-treated patients, four patients required invasive mechanical ventilation or ECMO on Day 1 of therapy, and four more cases subsequently required invasive mechanical ventilation (two cases on Day 6 and two cases on Day 9 of therapy). The 14-day, 28-day, and in-hospital mortality rates were 1.6%, 4.8%, and 7.9%, respectively. The major cause of death was superimposed infection.

The most common adverse event was diarrhea (54.0%), follow by nausea/vomit (7.9%), hepatitis (6.4%), and QT interval prolongation in EKG (6.4%). None of these adverse events were life-threatening.

### Factors Associated with Day-7 Clinical Improvement

To determine the factors associated with Day-7 clinical improvement, we compared patients with Day-7 clinical improvement (cases) with patients without Day-7 clinical improvement (controls). The characteristics of both groups are shown Table 1. The cases had a significantly lower age (47 vs. 59 years; *p* = 0.02), a significantly lower BMI (25.0 vs. 27.9; *p* = 0.04), a significantly lower baseline NEWS2 score (4 vs. 5; *p* = 0.003), and a significantly lower baseline six-point disease severity scale score (2 vs. 3; *p* < 0.001). Additionally, the baseline white blood cell count was significantly lower in the case group (5420 vs. 6810; *p* = 0.03). Although the median loading and maintenance doses of favipiravir were not statistically different between these groups, the proportion of patients in the control group who received a lower loading dose of favipiravir (≤45 MKD) trended higher compared with the case group (26.2% vs. 47.6%; *p* < 0.10).

Table 3 shows the results of multivariate analysis. A multivariate analysis revealed three factors that were negatively associated with Day-7 clinical improvement [odds ratio (95%CI); *p*-value]: older age [0.94 (0.89–0.99); *p* = 0.04], higher baseline NEWS2 score [0.64 (0.47– 0.88); *p* = 0.006], and a lower prescribed loading dose of favipiravir (≤45 MKD) [0.04 (0.005–0.4); *p* = 0.006].

**Table 3.**
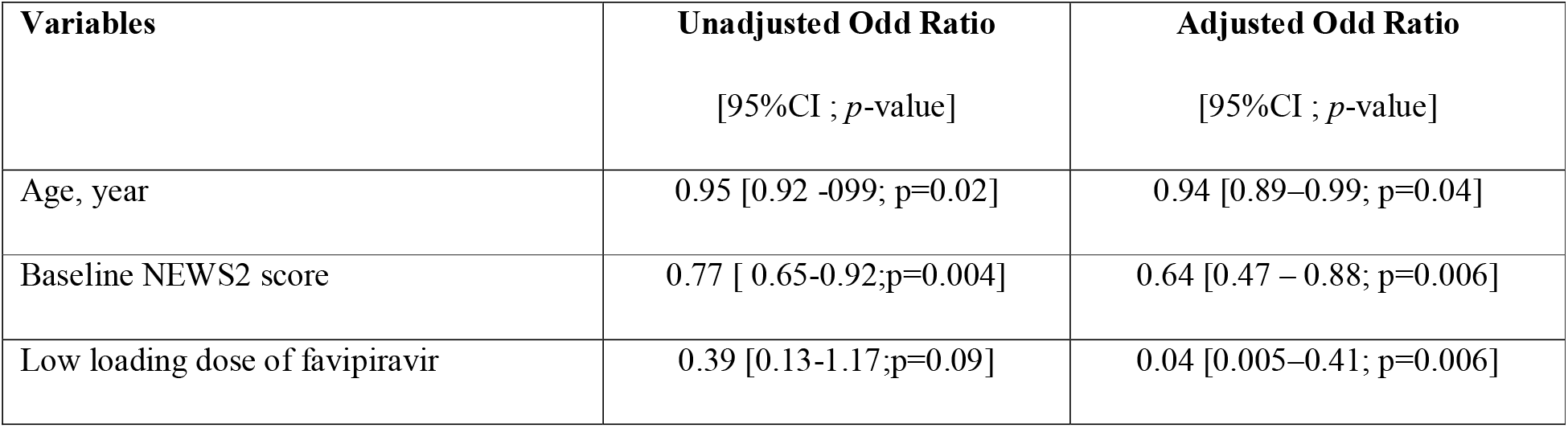
Factors associated with Day-7 clinical improvement

| **Variables** | **Unadjusted Odd Ratio**<br>[95%CI ; *p*-value] | **Adjusted Odd Ratio**<br>[95%CI ; *p*-value] |
| --- | --- | --- |
| Age, year | 0.95 [0.92 -0.99; *p*=0.02] | 0.94 [0.89–0.99; *p*=0.04] |
| Baseline NEWS2 score | 0.77 [ 0.65-0.92;*p*=0.004] | 0.64 [0.47 – 0.88; *p*=0.006] |
| Low loading dose of favipiravir | 0.39 [0.13-1.17;*p*=0.09] | 0.04 [0.005–0.41; *p*=0.006] |

## DISCUSSION

The Day-7 clinical improvement rate from our study was 67.7%, which is slightly lower than the Day-7 clinical recovery rate from the abovementioned RCT of favipiravir (71.4%).^8^ However, there were a few differences between these two studies. First, the definition of clinical recovery used in the abovementioned RCT was based mainly on clinical symptoms (e.g., fever, cough), whereas the definition of clinical improvement used in our study was based on improvement in oxygenation status. Our study included sicker patients with a higher proportion of patients who required mechanical ventilation (6.4%) as compared with the subjects of the unpublished RCT (0.9%). These differences may explain the slightly lower rate of favorable clinical responses observed in the present study.

Among the COVID-19 patients who did not require O_2_-supplementation, nearly all patients (92.6%) had clinical improvement within the first seven days of favipiravir therapy. However, only half of the patients who required O_2_-supplementation (47.2%) had clinical improvement within the first seven days of therapy. The rate of clinical improvement in these patients finally reached 75% on Day 14 and 83.3% on Day 28. Of the eight patients who required invasive mechanical ventilation or ECMO during their hospitalization, one patient died within the first 14 days. Therefore, the calculated 14-day mortality among this group was 12.5%. This number is similar to the 14-day mortality reported by a preliminary remdesivir RCT, in which 13 (10.4%) out of 125 patients who required mechanical ventilation or ECMO died.^12^ Based on these findings, the effectiveness of favipiravir for treating COVID-19 is promising, but this drug can be slow acting in more severe cases.

Our study identified older age and a higher baseline NEWS2 scale as poor prognostic factors for early clinical response. These findings are compatible with the results from many previous publications.^13-15^ We also explored other baseline variables (e.g., BMI, comorbidities); however the impact of those factors disappeared after the data were adjusted by the baseline NEWS2 scale.

Given that the optimal dose of favipiravir is still uncertain, we carefully explored the association between favipiravir dosage and patient outcome. Our study confirmed that a loading dose of favipiravir of ≤45 MKD was a poor prognostic factor for early clinical response. Therefore, a fixed favipiravir loading dose of 1600 mg twice daily for all patients with a BMI of <35 may be suboptimal for patients with a BMI of <35 but a body weight of ≥70 kg. Some might argue that this significant association may be a reflection of patients’ obesity, which was also known as a poor prognostic factor in COVID-19. However, our study did not find any association between the patient’s baseline BMI or body weight and the treatment outcome in the multivariate analysis.

Our study has several strengths. First, this study was a very early study to explore the effectiveness of favipiravir in active clinical cases of COVID-19. Second, this study included patients with differing disease severities; the patients ranged from mild pneumonia cases who did not require O_2_-supplementation to patients with life-threatening pneumonia who required mechanical ventilation or ECMO. This diverse subject pool allowed us to thoroughly explore the effectiveness of favipiravir and the clinical course of COVID-19 disease in various degrees of severity. Lastly, the daily NEWS2 scores and six-point disease severity scale scores were carefully collected and analyzed. Consequently, we can report nearly all important clinical outcomes and compare our findings with those of other clinical trials.^7, 8, 12^

Our study also has some limitations. First, the retrospective design resulted in a significant amount of missing data, especially for laboratory values. To resolve this issue, when performing the multivariate analysis, missing data was replaced by the mean value of a given variable. Second, majority of our patients also received chloroquine-based agent and protease inhibitors. Therefore, the good treatment response among our patients may be the synergistic results of triple combination of favipiravir, chloroquine-based agent and protease inhibitors. Furthermore, a high rate (54.0%) of diarrhea of the patients having favipiravir was probably related to protease inhibitor rather than favipiravir. Third, a sample size of 63 patients with COVID-19 pneumonia is not large enough to detect other associated factors with a low prevalence. Lastly, the generalizability of our findings may be an issue. Given that the study was conducted in tertiary care hospitals in Thailand, results may not be applicable to primary or secondary care settings or to COVID-19 patients in other countries.

In conclusion, our study reports the promising effectiveness of favipiravir for treating COVID-19 patients in a tertiary care hospital setting. No life-threatening adverse events were identified. In addition to older age and a high baseline NEWS2 score, a low loading dose of favipiravir (≤45 MKD) was also identified as a poor prognostic factor for early clinical improvement. Further studies to explore the optimal dose and the optimal timing of drug initiation for favipiravir should be performed.

## Data Availability

All datasets are available upon request.

## Notes

## Acknowledgments

The authors gratefully acknowledge the study team at all involved hospitals for their assistance of this study. Siriraj hospital: Mrs. Surangkana Samanloh; Taksin hospital: Mrs. Duangruethai Jankiew, Dr. Kittipong Sae-lao and Ms. Potjana Chularat; Vachira Phuket hospital: Ms. Somruedee Chatchawej and Dr. Busaya Santisant; Lerdsin hospital: Ms. Pranee Watagulsin; Central hospital: Ms. Nattasin Pimhom and Ms. Pornsikan Charutchocksawat. Additionally, we thank Katie Oakley, PhD, from Edanz Group (https://en-author-services.edanzgroup.com/) for editing a draft of this manuscript.

## Authors’ contributions

All authors made substantial contribution to the interpretation and analysis of data. The corresponding author had full access to the data and responsibility for the decision to submit the manuscript for publication.

## References

1. World Health Organization. Coronavirus disease (COVID-19) Situation Report – 139. https://www.who.int/docs/default-source/coronaviruse/situation-reports/20200607-covid-19-sitrep-139.pdf?sfvrsn=79dc6d08_2 (accessed June 8, 2020).

2. Srinivas P, Sacha G, Koval C. Antivirals for COVID-19. Cleve Clin J Med 2020.

3. Furuta Y, Komeno T, Nakamura T. Favipiravir (T-705), a broad spectrum inhibitor of viral RNA polymerase. Proc Jpn Acad Ser B Phys Biol Sci 2017; 93: 449–63.

4. Furuta Y, Takahashi K, Kuno-Maekawa M et al. Mechanism of action of T-705 against influenza virus. Antimicrob Agents Chemother 2005; 49: 981–6.

5. Bai CQ, Mu JS, Kargbo D et al. Clinical and Virological Characteristics of Ebola Virus Disease Patients Treated With Favipiravir (T-705)-Sierra Leone, 2014. Clin Infect Dis 2016; 63: 1288–94.

6. Du YX, Chen XP. Favipiravir: Pharmacokinetics and Concerns About Clinical Trials for 2019-nCoV Infection. Clin Pharmacol Ther 2020.

7. Cai Q, Yang M, Liu D et al. Experimental Treatment with Favipiravir for COVID-19: An Open-Label Control Study. Engineering (Beijing) 2020.

8. Chang Chen, Jianying Huang, Zhenshun Cheng et al. Favipiravir versus Arbidol for COVID-19: A Randomized Clinical Trial. medRxiv preprint https://doiorg/101101/2020031720037432 [e-pub ahead of print] 2020.

9. Wang Y, Fan G, Salam A et al. Comparative Effectiveness of Combined Favipiravir and Oseltamivir Therapy Versus Oseltamivir Monotherapy in Critically Ill Patients With Influenza Virus Infection. J Infect Dis 2020; 221: 1688–98.

10. Nguyen TH, Guedj J, Anglaret X et al. Favipiravir pharmacokinetics in Ebola-Infected patients of the JIKI trial reveals concentrations lower than targeted. PLoS Negl Trop Dis 2017; 11: e0005389.

11. Royal College of Physicians National Early Warning Score (NEWS) 2: Standardising the assessment of acute-illness severity in the NHS. Updated report of a working party London: RCP, 2017.

12. Beigel JH, Tomashek KM, Dodd LE et al. Remdesivir for the Treatment of Covid-19 - 353 Preliminary Report. N Engl J Med 2020.

13. Goyal P, Choi JJ, Pinheiro LC et al. Clinical Characteristics of Covid-19 in New York City. N Engl J Med 2020; 382: 2372-4.

14. Grasselli G, Zangrillo A, Zanella A et al. Baseline Characteristics and Outcomes of 1591 Patients Infected With SARS-CoV-2 Admitted to ICUs of the Lombardy Region, Italy. JAMA 2020.

15. Huang C, Wang Y, Li X et al. Clinical features of patients infected with 2019 novel coronavirus in Wuhan, China. Lancet 2020; 395: 497-506.

